# Safety and immunogenicity of SCB-2019, an adjuvanted, recombinant SARS-CoV-2 trimeric S-protein subunit COVID-19 vaccine in healthy 12–17 year-old adolescents

**DOI:** 10.1101/2023.02.22.23286317

**Authors:** Pio Lopez, Lulu Bravo, Erik Buntinx, Charissa Borja-Tabora, Hector Velasquez, Edith Johana Rodriguez, Camilo A. Rodriquez, Josefina Carlos, May Emmeline B. Montellano, Edison R. Alberto, Milagros Salvani-Bautista, Yung Huang, Branda Hu, Ping Li, Htay Htay Han, Carmen Baccarini, Igor Smolenov

## Abstract

We previously demonstrated efficacy of the COVID-19 vaccine candidate, SCB-2019, in adults in the SPECTRA phase 2/3 efficacy study. We extended the study to include 1278 healthy 12–17-year-old adolescents in Belgium, Colombia and the Philippines who received either two doses of SCB-2019 or placebo 21 days apart, to assess immunogenicity as neutralizing antibodies against prototype SARS-CoV-2 and variants of concern, and safety and reactogenicity as solicited and unsolicited adverse events with a comparator group of young adults (18–25 years). In participants with no evidence of prior SARS-CoV-2 infection SCB-2019 immunogenicity in adolescents was non-inferior to that in young adults; respective geometric mean neutralizing titers (GMT) against prototype SARS-CoV-2 14 days after the second vaccination were 271 IU/mL (95% CI: 211–348) and 144 IU/mL (116–178). Most adolescents (1077, 84.3%) had serologic evidence of prior SAR-CoV-2 exposure at baseline; in these seropositive adolescents neutralizing GMTs increased from 173 IU/mL (135–122) to 982 IU/mL (881–1094) after the second dose. Neutralizing titers against Delta and Omicron BA SARS-CoV-2 variants were also increased, most notably in those with prior exposure.

SCB-2019 vaccine was well tolerated with generally mild or moderate, transient solicited and unsolicited adverse events that were comparable in adolescent vaccine and placebo groups except for injection site pain – reported after 20% of SCB-2019 and 7.3% of placebo injections. SCB-2019 vaccine was highly immunogenic against SARS-CoV-2 prototype and variants in adolescents, especially in those with evidence of prior exposure, with comparable immunogenicity to young adults.

**Clinical trial registration:** EudraCT 2020-004272-17; ClinicalTrials.gov NCT04672395.

## INTRODUCTION

The global COVID-19 pandemic due to the SARS-CoV-2 virus resulted in over 600 million infections and over 6.5 million deaths [1]. Infections were initially most severe in older adults and those with underlying medical conditions, causing severe respiratory distress necessitating admission to intensive care units, often with fatal outcomes [2]. With this background initial vaccination efforts were targeted at these vulnerable groups, older adults and the chronically ill, then of the general adult population before expansion to adolescents. Therefore, clinical assessments of safety, tolerability, immunogenicity, and eventually efficacy of COVID-19 vaccines were performed in adult populations. The viral antigenic target used in most COVID-19 vaccines is the glycosylated Spike (S) protein, a trimeric protein consisting of two subunits, S1 and S2, which plays an essential role in viral binding, fusion and uptake into mammalian cells [3,4]. SCB-2019 is a recombinant SARS-CoV-2 S-protein produced in Chinese hamster ovary (CHO) cells fused in-frame to Trimer-Tag^©^ to preserve the native trimeric structure of S-protein in the prefusion form. Preclinical studies showed a vaccine formulation of SCB-2019 with the TLR9 agonist, CpG-1019 and aluminum hydroxide, induced protective immunity against SARS-CoV-2 *in vivo* in animal challenge studies [5]. Phase 1 studies in humans confirmed the safety, tolerability and immunogenicity of two doses of SCB-2019 vaccine administered 21 days apart to healthy adults [6].

Clinical development of SCB-2019 vaccine included a previously reported phase 2/3 study (SPECTRA) performed in adults 18 years of age and older at 31 sites in five countries; the phase 2 part studied safety, reactogenicity, and immunogenicity in subsets of participants, while the phase 3 part assessed efficacy against COVID-19 disease in all participants with no prior history of COVID-19 [7]. As national immunization efforts now include younger recipients, information about the impact of vaccination in this age group is an important resource so SPECTRA was extended to include adolescents, from 12–17 years of age. Here, we present the immunogenicity, reactogenicity and safety results from adolescents after a two-dose primary series of SCB-2019 vaccine or placebo administered 21 days apart, and compare the immunogenicity results with those previously observed in young adults, 18–25 years of age as part of the SPECTRA study.

## MATERIALS AND METHODS

SPECTRA is a double-blind, randomized, placebo-controlled, phase 2/3 study in 31 centres in five countries (Belgium, Brazil, Colombia, The Philippines, and South Africa) ongoing since March 24, 2021 [7]. The initial two co-primary objectives—reactogenicity of SCB-2019 vaccine in an embedded phase 2 study, and efficacy of SCB-2019 against COVID-19 in adults aged 18 years and older with no prior exposure to SARS-CoV-2 in the phase 3 study— have been reported previously [7].

Data presented here were obtained from an extension of the SPECTRA study to include 12– 17 year-olds enrolled in Belgium (1 site), Colombia (4 sites) and the Philippines (6 sites) from June 24, 2021 with a cut-off of June 14, 2022 for this analysis. Following a protocol amendment, the objectives of this extension study were to assess the safety and reactogenicity and neutralizing and binding antibody responses compared with placebo two weeks after the second of two doses of SCB-2019 vaccine administered 21 days apart, and a noninferiority analysis of immunity compared with a randomly selected set of young adults, 18–25 years of age, from SPECTRA. An exploratory analysis of efficacy was planned to measure the number of cases of COVID-19 occurring at least 14 days after the second vaccination in individuals with no evidence of prior SARS-CoV-2 infection. The study protocol and amendments were approved by all study center Institutional Review Boards and registered with ClinicalTrials.gov (identifier NCT04405908) and it was conducted according to International Conference of Harmonisation and Good Clinical Practice guidelines.

### Participants

Eligible participants were adolescents of either sex from 12 to 17 years of age inclusive at enrolment, who were healthy or had a stable pre-existing medical condition based on medical history and clinical assessment. All were screened for serum antibodies against SARS-CoV-2 as evidence of prior infection, and for acute exposure using a rapid COVID-19 Antigen test (RAT), which was repeated at each vaccination visit. Inclusion criteria included being able to understand and sign the informed consent and being available for the 12-month duration of the study. Female participants of childbearing potential were not to be pregnant or breastfeeding and had to agree to use protocol-approved forms of contraception from 30 days before the first vaccination to 90 days post second vaccination. Main exclusion criteria included any uncontrolled chronic medical disorders, any known or suspected impairment of the immune system due to known immunosuppressive conditions or any therapy with immunosuppressants or immunostimulants, known allergy to any vaccine components, malignancies, or prior receipt of any other SARS-CoV-2 vaccine.

### Vaccine

For SPECTRA SCB-2019 vaccine was supplied in three component parts: a sterile, clear to slightly opalescent and colorless solution for injection in a pre-filled syringe containing 720 µg SCB-2019 in 1·0 mL, the CpG-1018 adjuvant (Dynavax Technologies) in a 2·0 ml vial containing 12 mg/mL of a 22-mer phosphorothioate oligodeoxynucleotide in Tris buffered saline (24 mg per vial), both stored at 2°–8°C, and alum in vials of 10 mg/mL aluminium hydroxide (Alhydrogel^®^, Croda Health Care) kept at room temperature. The three components were mixed by gentle inversion at room temperature to a final vaccine formulation containing 30 µg SCB-2019 with 1·5 mg CpG-1018 and 0·75 mg alum per dose. Vaccine or placebo (0.5mL 0·9% sodium chloride for injection) were administered within 1 hour of preparation by intramuscular injection in the deltoid region using opacified syringes to maintain the blind for participants and study personnel responsible for the evaluation of any study endpoint.

### Procedures

Following screening, a Rapid COVID-19 Antigen test (RAT), and a full physical examination with an initial blood draw, eligible participants were vaccinated according to a centralized internet randomization system (IRS). Vaccine preparation and administration were performed by unblinded study personnel, but all participants and personnel involved in safety data collection and immunogenicity assessments were blinded to the study treatment.

On Days 1 and 22 each participant was vaccinated and monitored for 30 minutes post-vaccination for potential immediate post-vaccination reactions. Participants then completed daily electronic diary cards for 7 days to record solicited local reactions (pain, redness and swelling at the injection site), systemic adverse events (headache, fatigue, myalgia, arthralgia, loss of appetite, nausea, and chills) and body temperature. Solicited reactions and AEs were graded for severity (*see Supplementary Appendix pages 2–3*) by the participants. Any other adverse events were collected and assessed for causality at an investigator interview at the following study visit. Any unsolicited AEs were recorded up to Day 43. Serious adverse events (SAE), medically attended adverse events (MAAE), and adverse events of special interest (AESI) (*see Supplementary Appendix page 2*) were collected during the entire study period. SAEs and AESIs were to be reported immediately to the investigator and then to the study sponsor within 24 hours.

In addition to the RAT, all study participants had their Day 1 sera tested for anti-SARS-CoV-2 S-antigen by ELISA. A nasopharyngeal swab was collected from any participant suspected to be infected with SARS-CoV-2 virus for confirmation of COVID-19 by reverse transcription polymerase chain reaction (RT-PCR).

### Immunogenicity assessments

Sera prepared from blood draws on Days 1, 22 and 36 were stored at -80°C until shipment to the immunological laboratory (VisMederi srl, Siena, Italy) for analysis of randomly selected subsets of samples (the Immunogenicity Set). The primary immunogenicity endpoint was the anti-prototype SARS-CoV-2 neutralizing activity at Day 36 measured in a microneutralization assay (MN_50_) and expressed as geometric mean titers (GMT) with 95% confidence intervals (95% CI) and seroconversion rates (SCR). Titers against prototype SARS-CoV-2 were converted to International Units per mL (IU/mL) using the WHO standard 21/234. Cross-neutralizing activity was measured against the SARS-CoV-2 variants, Delta, Omicron BA.1, BA.2, BA.4 and BA.5. A secondary immunogenicity assessment was the anti-SCB-2019 IgG antibody titer, measured by ELISA and expressed as GMTs (95% CI) in IU/mL at each blood sampling timepoint. Details of the immunological assay methods are provided in the *Supplementary Appendix pages 4*.

### Statistics

In the amended protocol there was one statistical hypothesis for adolescents in the extension study: the non-inferiority of immune response in adolescents compared with young adults without evidence of prior SARS-CoV-2 infection assessed by prototype virus neutralization assay 14 days post dose 2. The population for the immunogenicity analysis was the per protocol immunogenicity set (PPS), consisting of those participants in the Immunogenicity Set with all blood samples available and analyzed from Days 1 and 36 and who had no evidence of prior SARS-CoV-2 exposure, i.e., no history of COVID-19 and negative RAT and serology test for SARS-CoV-2 at baseline. Immunogenicity was also measured in those in the immunogenicity subset with evidence of prior SARS-CoV-2 exposure, but participants who received other COVID-19 vaccine or had RT-PCR confirmed COVID-19 disease before Day 36 were excluded.

Non-inferiority (NI) of immunogenicity was assessed by GMT ratio and SCR difference. The GMT ratio (GMTR) was calculated via an ANCOVA model, including country as a covariate, along with the two-sided 95% CI for the GMR. The SCR was defined as the percentage of participants with post vaccination titer ≥ 4 x lower limit of quantitation (LLOQ) for initially seronegative participants, or at least a four-fold increase from baseline to post vaccination titer for initially seropositive participants. The SCR difference between adolescents and young adults was calculated with 2-sided 95% CI for the difference of SCR by the Miettinen and Nurminen method. NI was demonstrated if the lower limit (LL) of the 95% CI for the GMT ratio (GMT adolescents/GMT adults) exceeded 0.67 or if the SCR difference (SCR adolescents minus SCR adults) was greater than -10%.

The planned enrolment was 600 adolescents in the SCB-2019 group on the assumption that 10% would drop-out and 50–60% would not be eligible due to SARS-CoV-2 seropositivity, to provide a sample size of 240 evaluable adolescents. This would provide 93.7% power to demonstrate NI in comparison with 240 randomly selected young adults.

Other descriptive analyses calculated were GMT (95% CI) of antibody responses at each blood sampling timepoint for adolescents, and SCR and geometric mean fold rise (GMFR) at Days 22 and 36. Geometric mean values were calculated on log_10_ (titers/data) values, with subsequent antilog transformations applied, the 95% CI being calculated using normal distribution. The 95% CI for the SCR was computed by Clopper-Pearson method, and GMFR was calculated as the mean of the difference of logarithmically transformed assay results (post vaccine time point – baseline) and then anti-log transforming the mean. The associated 2-sided CIs were obtained by calculating CIs using paired t-distribution for the mean difference of the logarithmically transformed assay results and anti-log transforming back the confidence limits.

The Safety Analysis Set (SAS) consists of all participants who received at least one dose of their assigned vaccine or placebo. Reported summary statistics include counts and percentage of participants who reported at least one solicited local reaction, systemic adverse event or unsolicited adverse event (with severity and causality), and SAEs and AESIs, after the first and second doses. All analyses, and summaries were on group unblinded data performed using SAS^®^ software (version 9·4 or higher) or GraphPad Prism, v.6.0c.

## RESULTS

### Study population

Of 1350 adolescents screened, 70 were not considered to be eligible, and 1280 were enrolled and randomized equally to the two study groups (**Figure 1**). All but two participants, one each in the vaccine and placebo groups, received their assigned injections and made up the Safety Set. Median duration of participants in the study up to cut-off date was 97.0 days in both arms. Demographics of the two groups were generally similar (**Table 1**). Mean age was 14.3 years and most participants were enrolled in the Philippines (80.9% of each group), and there were slightly more male than female participants which was more evident in the vaccine group. Notably, 555 (86.9%) and 546 (85.4%) of the participants assigned to the SCB-2019 and placebo groups were already seropositive for S-protein at baseline and 4 (0.6%) and 3 (0.5%) had a history of prior COVID-19 infection (**Table 1**). Another 5 participants (3 SCB-2019, 2 placebo) had an RT-PCR-confirmed COVID-19 infection between Days 1 and 36.

**Table 1.** Demographics of the Safety Set and Per Protocol Immunogenicity Sets

|  | Safety Set |  | Per Protocol Immunogenicity sets |  |  |
| --- | --- | --- | --- | --- | --- |
|  | Adolescents |  | Adolescents |  | Young adults |
|  | SCB-2019<br>(N = 639) | Placebo<br>(N = 639) | SCB-2019<br>(N = 56) | Placebo<br>(N = 11) | SCB-2019<br>(N = 103) |
| **Characteristics** |  |  |  |  |  |
| **Sex – no. of participants (%)** |  |  |  |  |  |
| Male | 377 (59.0) | 344 (53.8) | 33 (59) | 6 (55) | 51 (49.5) |
| Female | 262 (41.0) | 295 (46.2) | 23 (41) | 5 (45) | 52 (50.5) |
| **Mean age ± SD, yr** | 14.3 ± 1.7 | 14.3 ± 1.7 | 14.4 ± 1.6 | 14.3 ± 2.2 | 21.2 ± 2.1 |
| **Hispanic or Latino ethnicity – no. of participants (%)** |  |  |  |  |  |
| Hispanic or Latino | 81 (12.7) | 76 (11.9) | 17 (30) | 5 (46) | 88 (85.4) |
| Not Hispanic or Latino | 558 (87.3) | 563 (88.1) | 39 (70) | 6 (54) | 15 (14.6) |
| **Race – no. of participants (%)** |  |  |  |  |  |
| American Indian /Alaskan native | 62 (9.7) | 60 (9.4) | 17 (30) | 5 (46) | 88 (85.4) |
| Asian | 518 (81.1) | 520 (81.4) | 9 (16) | 2 (18) | 1 (1.08) |
| Black or African American | 2 (0.3) | 1 (0.2) | 0 | 0 | 1 (1.0) |
| White | 57 (8.9) | 58 (9.1) | 30 (54) | 4 (36) | 13 (12.6) |
| **Country – no. of participants (%)** |  |  |  |  |  |
| Belgium (3 sites) | 58 (9.1) | 61 (9.5) | 31 (55) | 5 (46) | 15 (14.6) |
| Colombia (9 sites) | 64 (10.0) | 61 (9.5) | 17 (30) | 5 (46) | 88 (85.4) |
| The Philippines (10 sites) | 517 (80.9) | 517 (80.9) | 8 (14) | 1 (9) | 0 |
| **Baseline S-protein seropositive, n (%)** |  |  |  |  |  |
|  | 555 (86.9) | 546 (85.4) | 0 | 0 | 0 |
| **Known history of COVID-19, n (%)** |  |  |  |  |  |
|  | 4 (0.6) | 3 (0.5) | 0 | 0 | 0 |

**Figure 1:**
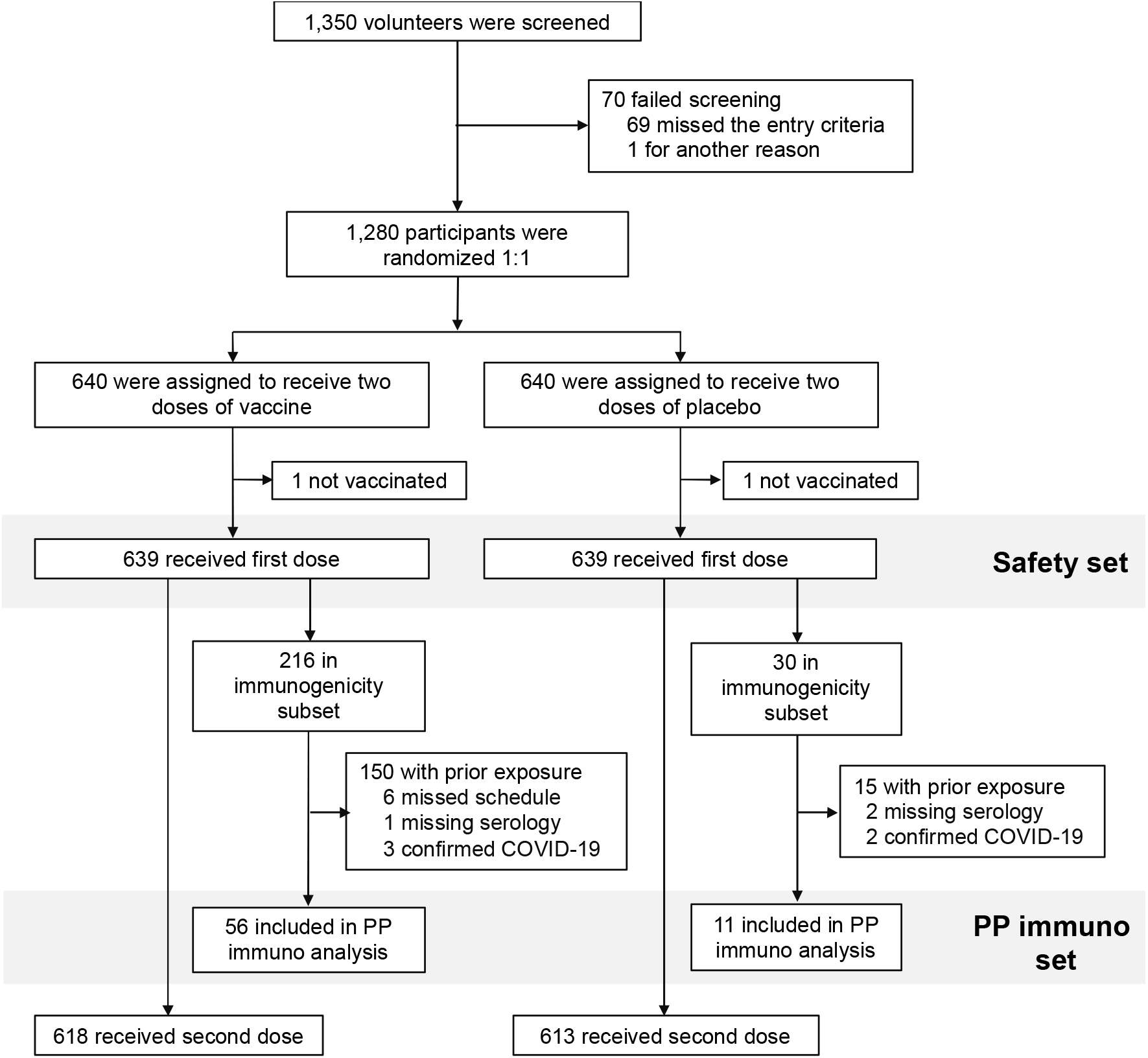
Study flow chart

The large proportion of enrolled participants who had evidence of prior SARS-CoV-2 exposure, approximately 87% of the study population, meant the number of eligible participants with no evidence of prior SARS-CoV-2 infection at baseline was too low for the efficacy assessment part of the study to have any conclusive result. A total of 103 baseline naïve young adults were included for the Per Protocol Set noninferiority analysis. Demographics of these 103 young adults were similar to those of the adolescents (**Table 1**).

### Immunogenicity -neutralizing antibodies

In the immunogenicity analysis in baseline SARS-CoV-2-naïve adolescents there was little response to SCB-2019 vaccination by Day 22, three weeks after the first dose, when only 1 of 55 (2%) vaccinees seroconverted for neutralizing antibodies and with little effect on the GMT (**Table 2**). However, by Day 36, two weeks after the second dose, there was a marked response in vaccinees and the pre-specified primary immunogenicity objective was met. At Day 36 GMTs were 271 IU/mL (95% CI: 211–348) in naïve adolescents and 144 IU/mL (116–178) in SARS-CoV-2-naïve young adults, a GMT ratio of 1.9 (95% CI: 1.3–3.0) which was above the pre-specified noninferiority criterion of 0.667 (**Table 2**). Similarly, respective Day 36 SCRs were 86% and 73%, a difference of 12.6% (95% CI: -1.3–24.3), again demonstrating non-inferiority. Placebo recipients did not display any neutralizing antibody responses.

**Table 2.** Geometric mean titers (GMT), seroconversion rate (SCR) and geometric mean-fold rise (GMFR) against prototype SARS-CoV-2 in the Per Protocol Adolescent Set

|  |  | Naïve Adolescents |  | Young adults |
| --- | --- | --- | --- | --- |
|  |  | SCB-2019 | Placebo | SCB-2019 |
| **Neutralizing antibodies (IU/mL) <sup>a</sup>** |  |  |  |  |
| **Day 1 (Baseline)** | N = | 55 | 9 |  |
|  | **GMT** | **13.7** | **12.5** |  |
|  | 95% CI) | (11.4, 16.6) | (-) |  |
| **Day 22** | N = | 56 | 10 |  |
|  | **GMT** | **15.6** | **12.5** |  |
|  | (95% CI) | (12.6, 19.3) | (-) |  |
|  | **SCR, n [%]** | **1 [2]** | **0** |  |
|  | (95% CI) | (0, 9.7) |  |  |
|  | **GMFR (95% CI)** | **1.1 (1.0, 1.3)** | **1.0** |  |
| **Day 36** | N = | 56 | 11 | 103 |
|  | **GMT** | **271** | **12.5** | **144** |
|  | (95% CI) | (211, 348) | (-) | (116, 178) |
|  | **GMT ratio for Adolescents vs Young adults = 1.9 (1.3, 3.0)** |  |  |  |
|  | **SCR, n [%]** | **47 [86]** | **0** | **75 [73]** |
|  | (95% CI) | (73, 94) |  | (63, 81) |
|  | **SCR difference for Adolescents vs Young adults = 12.6% (-1.3, 24.3)** |  |  |  |
|  | **GMFR (95% CI)** | **19.8 (15.0, 26.1)** | **1.0** |  |
| **SCB-2019 binding antibodies (IU/mL) <sup>b</sup>** |  |  |  |  |
| **Day 1 (Baseline)** | N = | 55 | 9 |  |
|  | **GMT** | **0.5** | **0.5** |  |
|  | (95% CI) | (0.5, 0.6) | (-) |  |
| **Day 22** | N = | 56 | 10 |  |
|  | **GMT** | **0.9** | **0.5** |  |
|  | (95% CI) | (0.7, 1.1) | (-) |  |
|  | **SCR, n [%]** | **3 [6]** | **0** |  |
|  | (95% CI) | (1.1, 15.1) |  |  |
|  | **GMFR (95% CI)** | **1.7 (1.3, 2.1)** | **1.0** |  |
| **Day 36** | N = | 55 | 11 |  |
|  | **GMT** | **14** | **0.5** |  |
|  | (95% CI) | (11.0, 17.8) | (-) |  |
|  | **SCR, n [%]** | **52 [95]** | **0** |  |
|  | (95% CI) | (84.9, 98.9) |  |  |
|  | **GMFR (95% CI)** | **26.5 (20.2, 34.7)** | **1.0** |  |

a) Neutralizing antibodies against prototype SARS-CoV-2 by microneutralization converted to International Units per mL (IU/mL) with the WHO international standard.
b) SCB-2019 binding antibodies against prototype SARS-CoV-2 virus by ELISA (IU/mL)

In contrast to the lack of response to the first dose in SARS-CoV-2-naïve adolescents, pre-exposed adolescents displayed an obvious booster effect to one dose. The first dose of SCB-2019 vaccine elicited a 4-fold increase in neutralizing GMT from 173 IU/mL (95% CI: 135– 222) at baseline to 702 IU/mL (619–796) with an SCR of 48% in these pre-exposed adolescents (**Table 3**). The GMT was further increased to 982 (881–1094) with an SCR of 60% after the second dose. Thus, SCB-2019 vaccine was able to boost the immune response initially elicited by SARS-CoV-2 exposure. In this pre-exposed group placebo recipients displayed a small decrease in GMT over the course of the study.

**Table 3.** Immunogenicity against prototype SARS-CoV in adolescents with and without (Naïve) serologic evidence of prior exposure to COVID-19 before vaccination as geometric mean titers (GMT), seroconversion rate (SCR) and geometric mean-fold rise (GMFR).

|  |  | Pre-exposed adolescents |  | Naïve adolescents with SCB-2019 <sup>a</sup> |
| --- | --- | --- | --- | --- |
|  |  | SCB-2019 | Placebo |  |
| **Neutralizing antibodies (IU/mL) <sup>b</sup>** |  |  |  |  |
| **Day 1**<br>(Baseline) | N = | 149 | 14 | 55 |
|  | **GMT** | **173** | **244** | **13.7** |
|  | 95% CI) | (135, 222) | (106, 559) | (11.4, 16.6) |
| **Day 22** | N = | 148 | 15 | 56 |
|  | **GMT** | **702** | **225** | **15.6** |
|  | (95% CI) | (619, 796) | (114, 443) | (12.6, 19.3) |
|  | **SCR, n [%]** | **71 [48]** | **1 [7]** | **1 [2]** |
|  | (95% CI) | (39.7, 56.3) | (0.2, 33.9) | (0, 9.7) |
|  | **GMFR (95% CI)** | **4.0 (3.2, 5.0)** | **1.0 (0.6, 1.7)** | **1.1 (1.0, 1.3)** |
| **Day 36** | N = | 149 | 15 | 55 |
|  | **GMT** | **982** | **166** | **271** |
|  | (95% CI) | (881, 1094) | (82.9, 333) | (211, 348) |
|  | **SCR, n [%]** | **90 [60]** | **1 [7]** | **47 [86]** |
|  | (95% CI) | (52.1, 68.3) | (0.2, 33.9) | (73, 94) |
|  | **GMFR (95% CI)** | **5.7 (4.5, 7.1)** | **0.7 (0.4, 1.2)** | **19.8 (15.0, 26.1)** |
| **SCB-2019 binding antibodies (IU/mL) <sup>c</sup>** |  |  |  |  |
| **Day 1**<br>(Baseline) | N = | 149 | 14 | 55 |
|  | **GMT** | **5.7** | **7.5** | **0.5** |
|  | (95% CI) | (4.5, 7.2) | (3.8, 14.8) | (0.5, 0.6) |
| **Day 22** | N = | 147 | 15 | 56 |
|  | **GMT** | **27.4** | **5.8** | **0.9** |
|  | (95% CI) | (23.3, 32.3) | (3.2, 10.5) | (0.7, 1.1) |
|  | **SCR, n [%]** | **72 [49]** | **1 [7]** | **3 [6]** |
|  | (95% CI) | (40.7, 57.3) | (0.2, 33.9) | (1.1, 15.1) |
|  | **GMFR (95% CI)** | **4.7 (3.6, 6.1)** | **0.8 (0.5, 1.3)** | **1.7 (1.3, 2.1)** |
| **Day 36** | N = | 149 | 15 | 55 |
|  | **GMT** | **31.3** | **4.1** | **14** |
|  | (95% CI) | (26.4, 37.0) | (2.7, 6.4) | (11., 0, 17.8) |
|  | **SCR, n [%]** | **84 [56]** | **1 [7]** | **52 [95]** |
|  | (95% CI) | (48.0, 64.5) | (0.2, 33.9) | (84.9, 98.9) |
|  | **GMFR (95% CI)** | **5.5 (4.3, 7.2)** | **0.6 (0.3, 0.9)** | **26.5 (20.2, 34.7)** |

a) Naïve adolescents had no evidence of prior exposure to SARS-CoV-2 or COVID-19
b) Neutralizing antibodies against prototype SARS-CoV-2 virus determined by microneutralization converted to International Units per mL (IU/mL) with the WHO international standard.
c) SCB-2019 binding antibodies against prototype SARS-CoV-2 virus determined by ELISA (IU/mL)

Cross-neutralizing responses against Delta and Omicron BA subvariants were measured in those with and without evidence of prior SARS-CoV-2 exposure (**figure 2**). In SARS-CoV-2-naïve participants with no baseline cross-neutralizing activity against any of the SARS-CoV-2 strains tested, there were marked responses against Delta and Omicron BA.2 and a smaller response against Omicron BA.5, but little effect against Omicron BA.1 and BA.4. In those with some cross-neutralizing activity at baseline there were marked responses achieving an approximate 10-fold increase in GMT against each strain.

**Figure 2:**
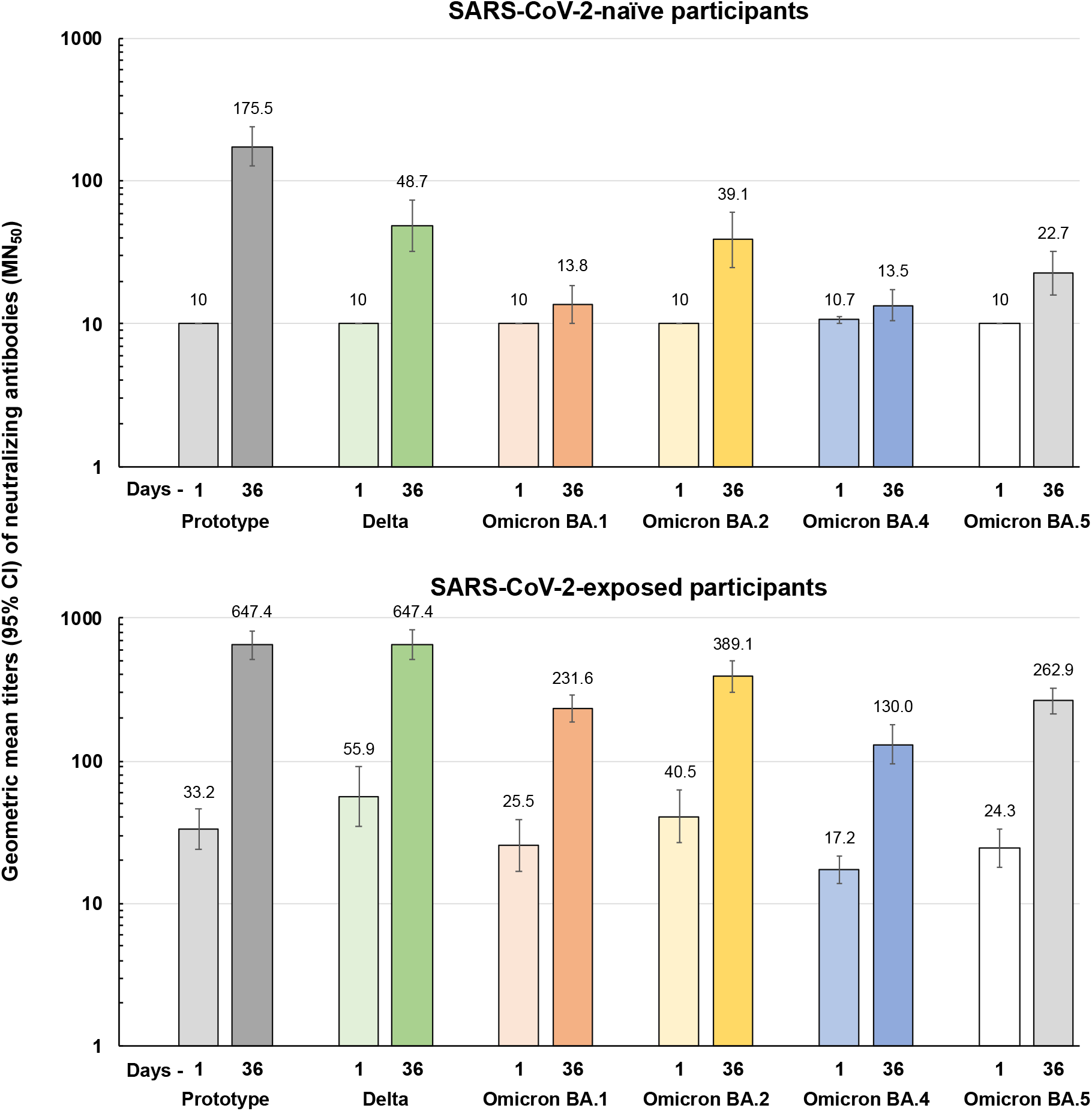
Geometric mean titers (95% CI bars) of neutralizing antibodies against prototype SARS-CoV-2 virus and the Delta and indicated Omicron BA variants on Days 1 and 36 in adolescents according to their baseline serostatus for known exposure to SARS-CoV-2. Numbers above columns show GMTs.

### Immunogenicity – SCB-2019-binding antibodies

The same pattern of responses was observed when the immunogenicity was measured as SCB-2019-binding antibodies by ELISA. In naïve adolescents only 6% (3/55) seroconverted after the first dose with a small increase in GMT from 0.5 IU/mL (95% CI: 0.5–0.6) to 0.9 (0.7–1.1). However, after the second dose 95% (52/55) seroconverted, achieving a GMT of 14 IU/mL (11.0 –17.8), a GMFR of 26.5, while placebo recipients did not display any response after either dose. In pre-exposed adolescents the first dose elicited a 4.7-fold increase in GMT of SCB-2019-binding antibodies, from 5.7 IU/mL (4.5–7.2) to 27.4 (23.3– 32.3) with a smaller increase after the second dose to 31.3 IU/mL (26.4–37.0). The high level of SCB-2019-binding antibodies at baseline indicates significant cross-reactivity between SCB-2019 and natural immunity due to exposure to wild-type SARS-CoV-2.

### Safety assessments

There were no deaths during the study and of the six SAEs reported, two in the SCB-2019 group (one case of constipation and one fractured clavicle) and four (cases of appendicitis, bronchitis, and nervous syncope, and a participant who sustained ankle and forearm fractures) in the placebo group, none were considered by the investigator to be related to study interventions (**Table 4)**. Medically attended adverse events and AESIs were balanced between the two groups.

**Table 4.** Unsolicited adverse events, serious adverse events and MAAE in the two study groups from Day 1 to Unblinding.

|  |  | **SCB-2019** | **Placebo** |
| --- | --- | --- | --- |
|  | **N =** | **639** | **639** |
| **Unsolicited adverse events (Day 1 to Unblinding)** | n Participants (n events) | 93 (114) | 86 (114) |
|  | [% of participants] | [14.6%] | [13.5%] |
|  | Participants with a related event (n events) | 7 (7) | 6 (7) |
|  | Severe events [%] | 0 | 1 (0.2) |
| **Serious adverse events (SAE)** | Any, n (%) | 2 (0.3) | 4 (0.6) |
|  | Related | 0 | 0 |
| **Medically attended AE (MAAE)** | n (%) | 55 (8.6) | 49 (7.7) |
| **AE of special interest (AESI)** | n (%) | 6 (0.9) | 4 (0.6) |
| **AE leading to early termination from study** | n (%) | 0 | 0 |
| **Death** | n (%) | 0 | 0 |

The incidences of solicited local reactions shown in **Figure 3**, consisting almost exclusively of injection site pain, were higher in adolescents who received SCB-2019 vaccine (∼20%) than placebo recipients (∼7.3%), but were comparatively lower than in young adults who received SCB-2019 vaccine (∼44%). Local reaction rates in both adolescent groups were lower after the second dose, but not in the young adults. The severity was similar across groups and all reactions resolved within a few days. For solicited systemic adverse events frequencies and severities were similar after the first dose in adolescents who received SCB-2019 vaccine or placebo (**Figure 4**), and again were lower than observed in the young adults. The most frequent events were headache and fatigue. The incidences of the solicited systemic AEs were lower after the second dose in both adolescents and the young adults, but the latter still reported higher rates of solicited AEs.

**Figure 3:**
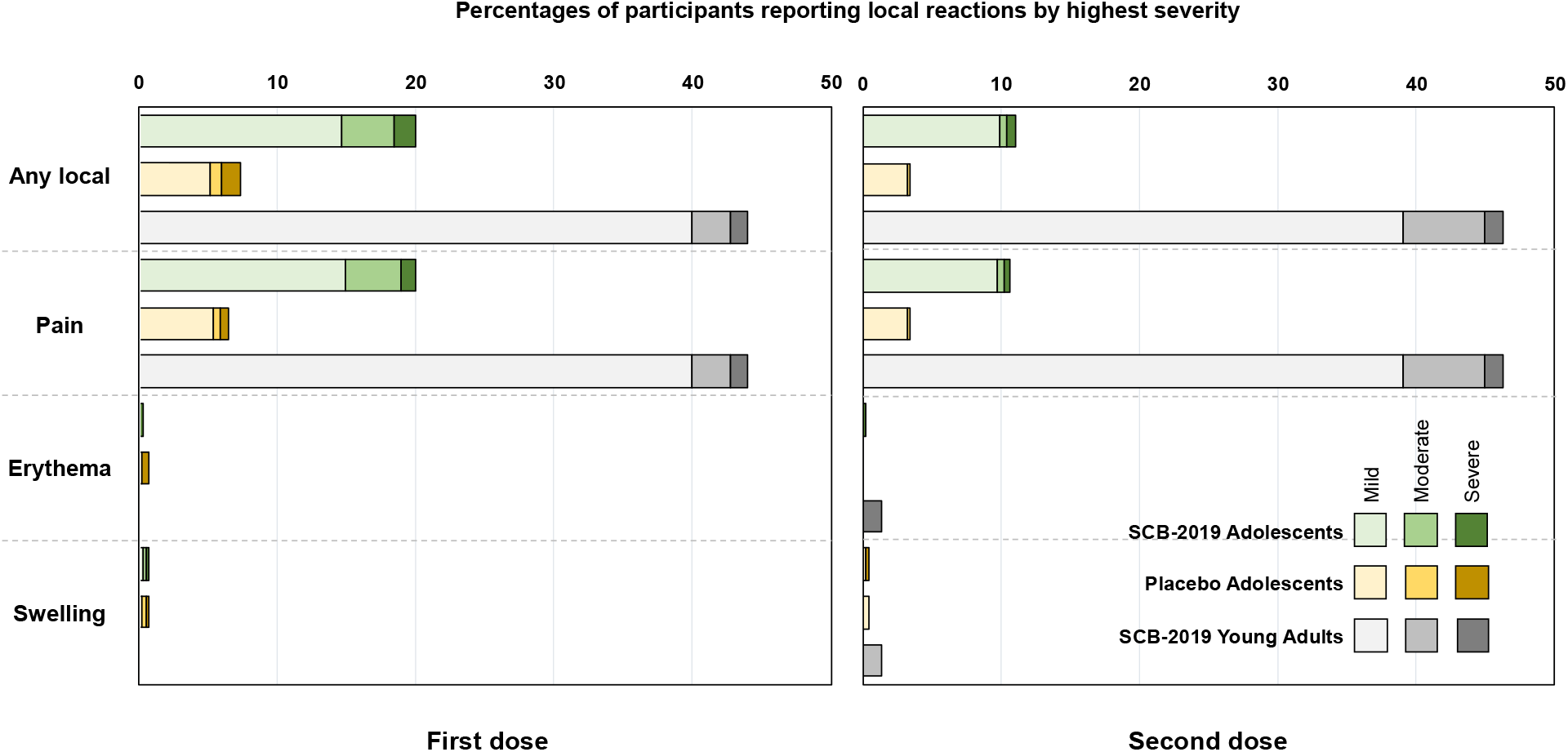
Solicited local reactions the 7 days after the first and second vaccinations by highest severity in the two adolescent study groups and young adults (n= 75) from the SPECTRA study.

**Figure 4:**
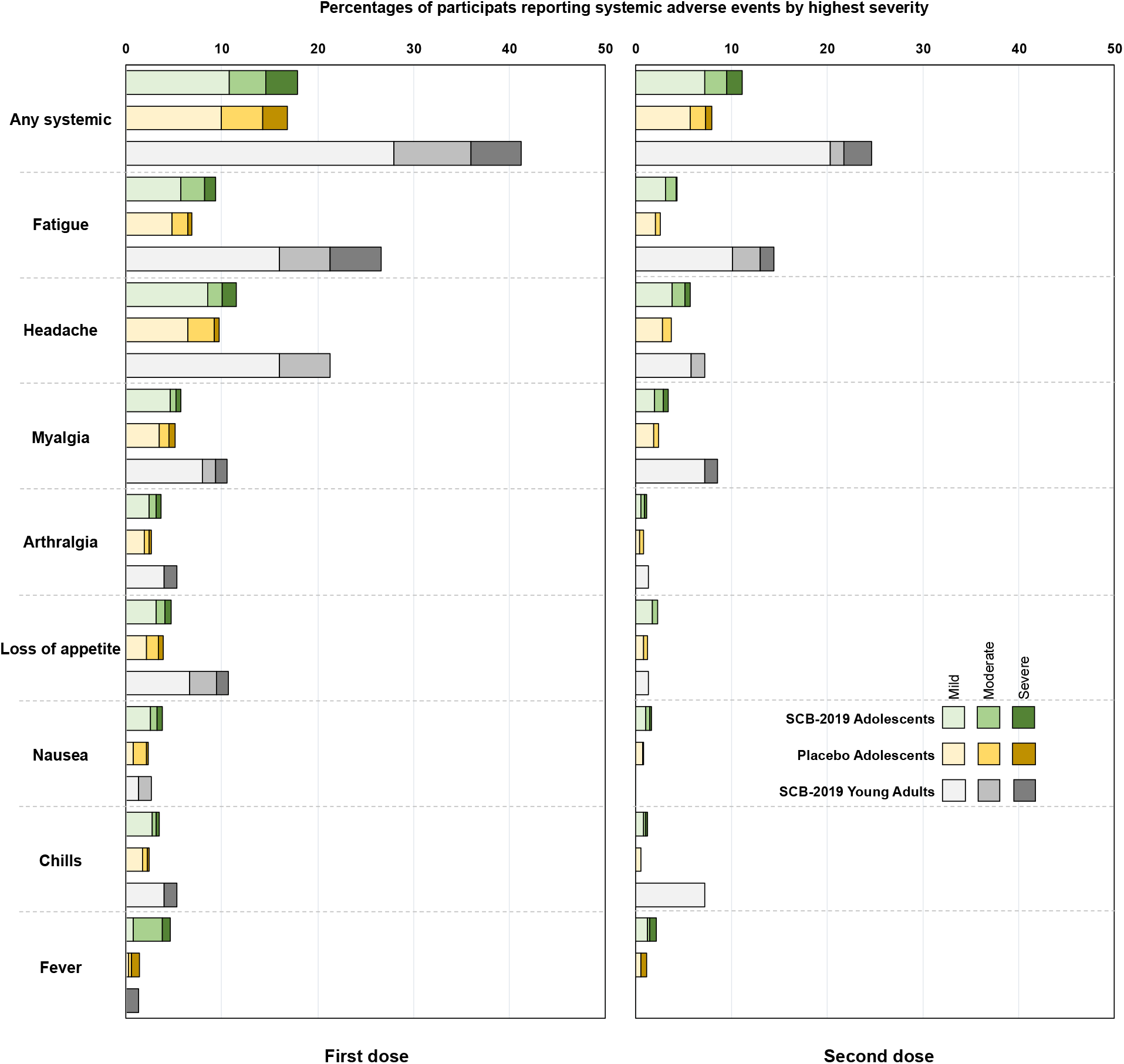
Solicited systemic adverse events in the 7 days after the first and second vaccinations by highest severity in the two adolescent study groups and young adults (n= 75) from the SPECTRA study.

Unsolicited adverse events, occurring up to unblinding were balanced between the two groups of adolescents (**Table 4**). Most were considered to be unrelated to study treatments, and only one was considered severe, a fractured forearm in a placebo recipient. The investigator did not consider any of the of the six reported SAEs, 2 in the SCB-2019 group and 4 in the placebo group, to be related to the vaccinations.

## DISCUSSION

The primary objective of this extension of the SPECTRA study was to assess the immunogenicity of two doses of SCB-2019 vaccine in adolescents aged from 12 to 17 years, including a noninferiority comparison with immunogenicity observed in a sample population of young adults aged 18 to 25 years from the SPECTRA study. We confirmed the non-inferiority of immune responses to two doses of SCB-2019 vaccine in previously SARS-CoV-2 naïve adolescents in whom the GMT of neutralizing antibodies against prototype

SARS-CoV-2 two weeks after the second dose was approximately two-fold higher than the response observed in the young adults, and the seroconversion rate was also higher. We also confirmed the excellent tolerability of SCB-2019 vaccination in adolescents, who presented with lower rates of solicited local and systemic adverse events than the young adults, and had no vaccine-related serious adverse events.

It was not possible to do a meaningful assessment of the original efficacy objective in this study as 1101 of 1278 (86%) adolescents had evidence of prior exposure to SARS-CoV-2 before vaccination. However, the neutralizing responses measured in adolescents compare well with those measured in adults from 18 to approximately 80 years of age in the phase 2/3 efficacy trial of SCB-2019 vaccine in which efficacies of 67.2%, 83.7% and 100% against any severity, moderate-to-severe and severe COVID-19, respectively, were observed [7]. In that adult population the neutralizing GMTs at Day 36 were 224 IU/mL (95% CI: 194.0– 258.7; n = 220) and 1831 IU/mL (1546–2170; n = 118) in SARS-CoV-2-naïve and SARS-CoV-2-exposed participants, respectively [8]. The 271 IU/mL (211–348) and 982 IU/mL (881–1094) in SARS-CoV-2-naïve and SARS-CoV-2-exposed adolescents measured in the present study are consistent with those values suggesting similar efficacy would have been achieved.

In adolescents with serological evidence of prior SARS-CoV-2 exposure, there was a four-fold increase in neutralizing antibodies after one dose of SCB-2019 vaccine, whereas in SARS-CoV-2-naïve adolescents the first dose of SCB-2019 vaccine had little effect on the immunogenicity consistent with previous observations [6,8]. The magnitude of the response in pre-exposed adolescents after the first dose of SCB-2019 vaccine suggests there is a booster response on top of the immunity induced by natural exposure to SARS-CoV-2 in these individuals. A second dose in those participants elicited a small further increase in GMT. Thus, administration of two doses of SCB-2019 vaccine was safe and immunogenic in adolescents irrespective of their serological status against SARS-CoV-2 before vaccination. Further, there were also marked increases in cross-neutralizing activity against all tested SARS-CoV-2 variants in the majority of participants who displayed serologic evidence of prior exposure to the virus, although there was less effect on those who were SARS-CoV-2-naïve.

The observation of a marked immune response, including cross-neutralizing activity against SARS-CoV-2 variants, in initially seropositive adolescents is serendipitous as it confirms that SCB-2019 vaccine is suitable for use as booster vaccination in those who may previously have had a symptomatic or asymptomatic COVID-19 infection. In the current situation of the COVID-19 pandemic the most likely use for SCB-2019 vaccine is as a booster vaccination as most of the population now will either have already been infected by SARS-CoV-2, as illustrated by our baseline data, or have been vaccinated. The potential of SCB-2019 as a heterologous booster vaccine has already been investigated in Brazilian adults previously vaccinated with two doses of the adenovirus-vector vaccine, ChAdOx1-S [9]. That study found a heterologous dose of SCB-2019 vaccine induced significantly higher titers of neutralizing antibodies than a homologous ChAdOx1-S booster dose, against the prototype wild-type virus and notably also against the Beta, Delta, Gamma and Omicron variants.

As increasing proportions of populations are either at least partially vaccinated against SARS-CoV-2 or have recovered from COVID-19 infections, the prevention of future outbreaks due to emerging variants will rely heavily on maintaining a high level of immunity through booster vaccination campaigns. Increasing experience indicates that the most success with such campaigns will be through using heterologous booster vaccines [10–13]. Following the previous experience of the global pandemic, such campaigns are only likely to be successful if they include adolescents as well as the elderly and adults, so our demonstration of the safety and immunogenicity of SCB-2019 vaccine in adolescents is important to inform planning of such campaigns. Other COVID-19 vaccines have been shown to be well tolerated and immunogenic in the age groups similar to ours [14–17], although there have been reports of rare cases of myocarditis and pericarditis in male adolescents following mRNA COVID-19 vaccines [18] and autoimmune diseases in children and adolescents [19].

This study was limited in that only a small proportion of the vaccinated adolescents had no evidence of prior SARS-CoV-2 exposure, such that the intended efficacy assessment was not viable. However, this is a consequence of the rapid spread of the COVID-19 pandemic and the current global situation where the peak of the pandemic has subsided except in certain localized outbreaks. The demonstration of good tolerance and high immunogenicity in pre-exposed adolescents is therefore important as it allows the use of SCB-2019 as a booster vaccine without requiring pre-screening for the vaccinee’s history of SARS-CoV-2 exposure. The study confirms that two doses of SCB-2019 vaccine induce neutralizing antibodies against the prototype wild-type SARS-CoV-2 virus in adolescents. The noninferiority of the adolescent response to that observed in young adults in whom efficacy was demonstrated in the SPECTRA study suggests that adolescents will also be protected. Further, in the majority of the population who now have some serologic evidence of prior exposure SCB-2019 vaccination induces cross-neutralizing activity against circulating Variants of Concern, particularly the various sub-lineages of Omicron which currently predominate. This supports the suitability of SCB-2019 vaccine for the currently intended use of COVID-19 vaccines, as booster doses in vaccine-primed and/or exposed populations.

In conclusion, we have demonstrated that SCB-2019 vaccine is well tolerated and highly immunogenic in adolescents for 12 to 17 years of age, irrespective of their previous experience with SARS-CoV-2 and can be safely administered to this group as a booster vaccine to maintain a high level of immunity to prevent future outbreaks.

## Data Availability

The datasets, including the redacted study protocol, redacted statistical analysis plan, and individual participants data supporting the results reported in this article, will be available three months from initial request, to researchers who provide a methodologically sound proposal. The data will be provided after its de-identification, in compliance with applicable privacy laws, data protection and requirements for consent and anonymization.

## ACKNOWLEDGEMENTS

We are grateful to all the volunteers and the study staff at the different study centers. We thank the he personnel at Vismederi SRL (Siena, Italy) for their expertise in performing the immunogenicity assays, and Dr. Keith Veitch (keithveitch communications, Amsterdam, the Netherlands) for drafting and editorial management of the manuscript.

## CONFLICTS OF INTEREST

YH, BH, PL, HHH, CB and IS are full-time employees of Clover Biopharmaceuticals. Other authors have no interest to declare.

## FINANCIAL DECLARATION

This work was supported by grants from Coalition for Epidemic Preparedness Innovations (CEPI), grant number PRJ-6052.

## DISCLAIMER

Any opinions, findings, and conclusions expressed in this material are those of the authors.

## CONTRIBUTORS

IS, CB, PL, BH and HHH designed the study, medical oversight was provided by YH and CB, and data analysis support by PL. Interpretation and writing of the manuscript was by all authors led by IS and a medical writer. All authors reviewed the manuscript and approved the submission for publication.

## Notes

### Clinical Trial

NCT04405908

### Author Declarations

Ethics committee of European Medicines Agency gave ethical approval for this work Ethics committee of China Center for Drug Evaluation gave ethical approval for this work Ethics committee of Anvisa Brazil gave ethical approval for this work Ethics committee of the Philippines Food and Drug Administration gave ethical approval for this work

