## Supplementary appendix for "Safety and immunogenicity of SCB-2019, an adjuvanted, recombinant SARS-CoV-2 trimeric S-protein subunit COVID-19 vaccine in healthy 12–17 year-old adolescents"

Smolenov I, et al.

**Supplementary Appendix**

Definition of a Serious Adverse Event (SAE)	page 2.

Definition of an Adverse Event of Special Interest (AESI)	page 2.

Severity grading for solicited local reactions and systemic adverse events	page 2.

IMMUNOLOGICAL METHODS

SARS-CoV-2 wild-type microneutralization	page 4.

SCB-2019 binding ELISA	page 4.

**Definition of a Serious Adverse Event (SAE)**

An SAE is any adverse event that:

- Results in death
- Is life-threatening
- Requires inpatient hospitalization or prolongation of existing hospitalization
- Results in persistent disability/incapacity
- Is a medically important event in the opinion of the investigator
- Is a congenital anomaly/birth defect

**Definition of an Adverse Event of Special Interest (AESI)**

An AESI is a potential immune-mediated disease including autoimmune disease and other inflammatory and/or neurologic disorder of interest which may or may not have an autoimmune etiology. Additionally, adverse events potentially associated with COVID-19 as defined by the Safety Platform for Emergency vACcines (SPEAC),* were also reported as AESIs in this study.

* https://brightoncollaboration.us/wpcontent/uploads/2020/11/SPEAC_SO1_2.2_2.3-SO2-D2.0_Addendum_AESI-Priority-TiersAug2020-v1.2.pdf

| **Severity grading for solicited local reactions and systemic adverse events** | | | |
| --- | --- | --- | --- |
|  | Mild | Moderate | Severe |
| Pain | No interference with daily activities | Interferes with daily activities | Prevents daily activity |
| Erythema | 25-50 mm | 51-100 mm | >100 mm |
| Swelling | 25-50 mm | 51-100 mm | >100 mm |
| Fatigue | No interference with daily activities | Interferes with daily activities | Prevents daily activity |
| Headache | No interference with daily activities | Interferes with daily activities | Prevents daily activity |
| Myalgia | No interference with daily activities | Interferes with daily activities | Prevents daily activity |
| Arthralgia | No interference with daily activities | Interferes with daily activities | Prevents daily activity |
| Loss of appetite | Eating less than usual/no effect on normal activity | Eating less than usual/interfered with normal activity | Not eating at all |
| Nausea | No interference with daily activities | Interferes with daily activities | Prevents daily activity |
| Chills | No interference with daily activities | Interferes with daily activities | Prevents daily activity |
| Fever | 38.0º–38.4ºC | 38.5º–38.9ºC | >39.0ºC |

**IMMUNOLOGICAL METHODS**

***SARS-CoV-2 wild-type microneutralization assay (WT-MN)***

Serum samples were heat inactivated for 30 minutes at 56⁰C and eleven two-fold serial dilutions of test samples were prepared in a separate dilution plate. Sera were mixed with an equal volume of SARS-CoV-2, hCoV-19/Australia/VIC01/2020 (GenBank MT007544.1), and incubated for 1 hr at 37°C, 5% CO_2_. The virus/serum mixtures (200 TCID_50_ units/well) were then transferred in duplicate to sub-confluent Vero E6 cell monolayer plates, pre-seeded 24 hours beforehand in 96 well plates at 1·5 x 10^4^ cells/well. Plates were incubated for 3 days at 37°C, 5% CO_2_. The residual non-neutralized virus was detected via cytopathic effect (CPE) by microscopic scoring. The neutralization titer was expressed as the reciprocal of the highest dilution at which 50% of the replicate wells were protected from infection (MN_50_) and converted to International units per mL using the WHO international standard for SARS-CoV-2 antibodies.

***SCB-2019 binding ELISA***

Maxisorp plates were coated with 1μg/ml SCB-2019 at 4°C overnight and blocked with 2% non-fat milk in PBS containing 0·05% Tween-20 (PBST). Eight two-fold serial dilutions of serum samples starting from a 1:25 initial dilution were added to the blocked and washed SCB-2019-coated plates and incubated for 1h at 37°C. Plates were then washed, incubated with HRP-conjugated anti-human IgG for 1 hr at 37°C, washed again and the colorimetric signals were developed using TMB substrate for 3 min before stopping the reaction with 1N sulfuric acid. Optical density (OD) was measured at 450/650 nm. The EC_50_ of each test sample was calculated using a non-linear four parameter regression curve using GraphPad Prism, v.6.0c.
